# An enhanced method for calculating trends in infections caused by pathogens transmitted commonly through food

**DOI:** 10.1101/2022.09.14.22279742

**Authors:** Daniel L. Weller, Logan C. Ray, Daniel C. Payne, Patricia M. Griffin, Robert M. Hoekstra, Erica Billig Rose, Beau B. Bruce

## Abstract

This brief methods paper is being published concomitantly with “Preliminary Incidence and Trends of Infections Caused by Pathogens Transmitted Commonly Through Food— Foodborne Diseases Active Surveillance Network, 10 U.S. Sites, 2016–2021” in Morbidity and Mortality Weekly Reports (MMWR). That article describes the application of the new model described here to analyze trends and evaluate progress towards the prevention of infection from enteric pathogens in the United States.

---

In 2022, we implemented a new model for estimating trends in illnesses in the ten U.S. sites conducting population-based surveillance for laboratory-confirmed enteric illnesses as part of the Foodborne Diseases Active Surveillance Network (FoodNet). This model was developed to address some limitations of the model used in earlier annual FoodNet reports. Principal improvements include the use of spline regression, an interaction term between site and year, and the treatment of year as continuous (the previous model treated year as a categorical variable). The new method reduces sensitivity to single-year aberrations (e.g., outbreaks), accounts for site-specific trends, and reduces biases toward more populous sites (Figures 1 & 2).

**Figure 1:**
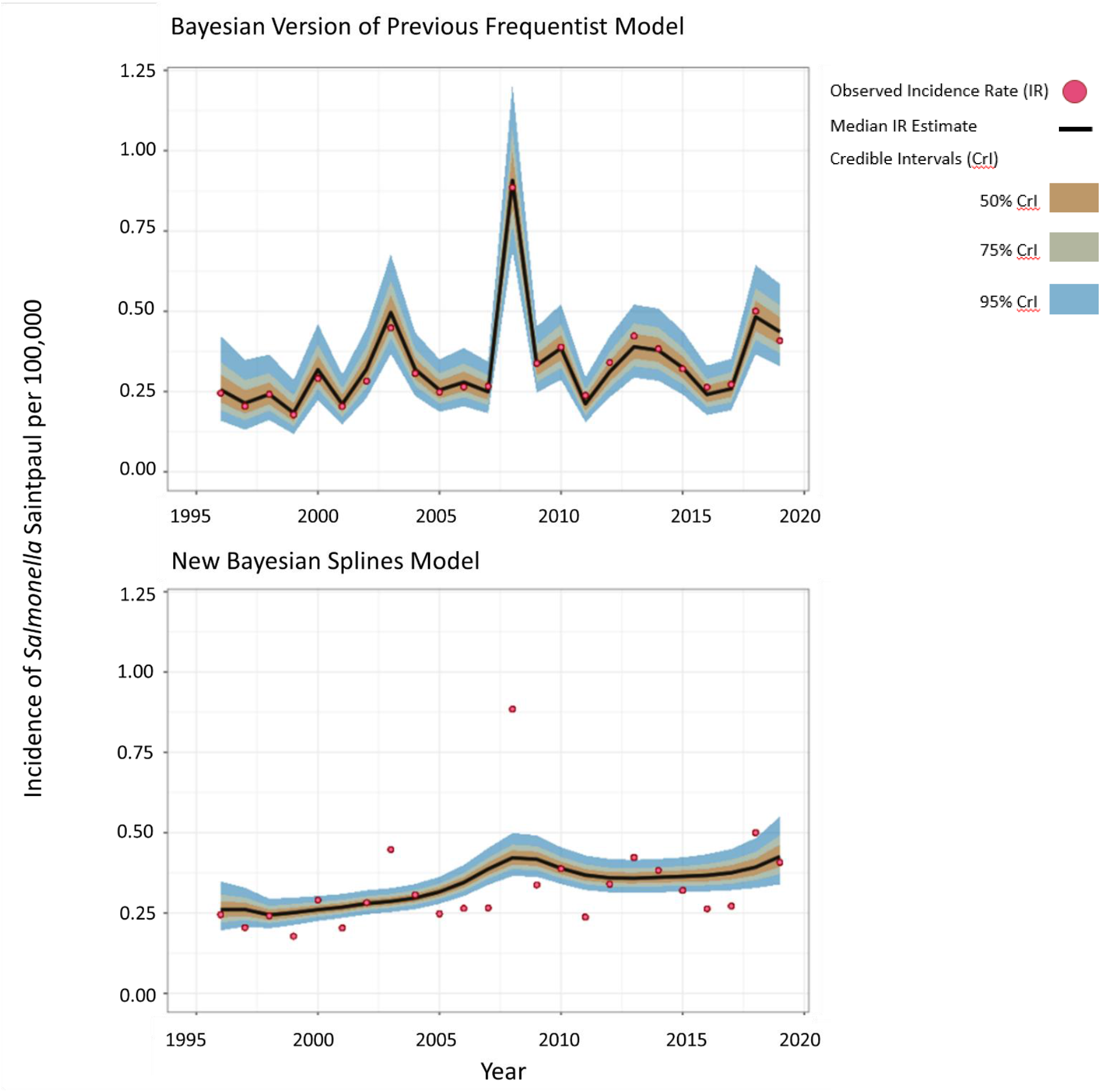
Actual (dots) and median estimates (line) of *Salmonella* serotype Saintpaul illnesses per 100,000 population in the FoodNet catchment during 1996 to 2019 according to the previous model^a^ (top) and the new Bayesian splines model (bottom). The shading represents the 50%, 75%, and 95% credibility intervals (CrI). Note how sensitive the previous model is to single-year aberrations, like the 2008 jalapeño and serrano pepper outbreak (vertical blue line), compared with the new model. ^a^The model used in prior MMWRs was frequentist; a Bayesian version of that model was created to make outputs that could be compared with those of the new model.

**Figure 2:**
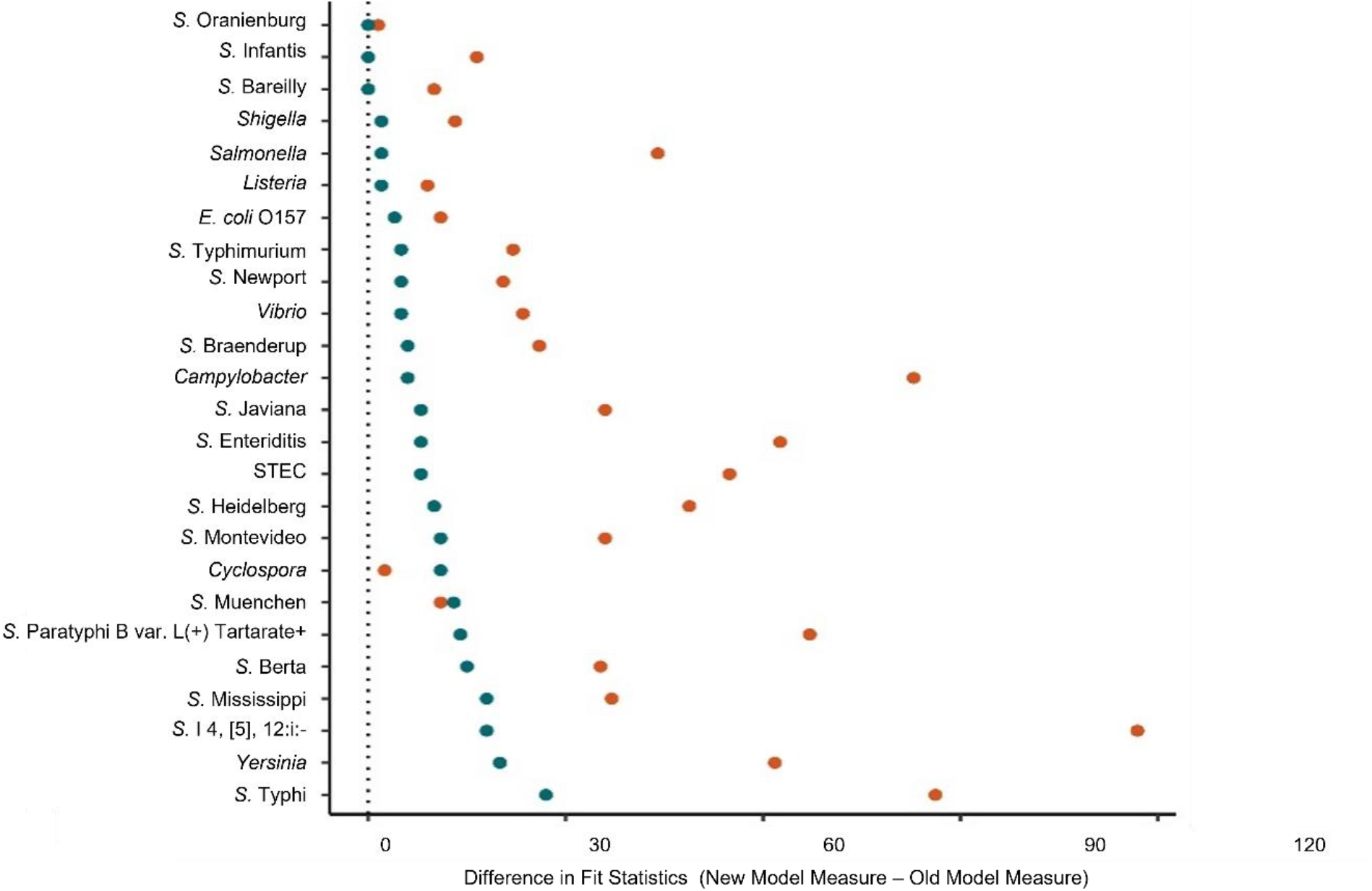
Improved fit of the new Bayesian splines model compared with a Bayesian version of the previous frequentist model. Fit was quantified using two statistical measures: estimated expected log pointwise predictive density (ELPD; orange) and R^2^* (blue). The difference was calculated between the new and old models for each measure. Results are shown for all pathogens tracked by FoodNet, including Shiga toxin-producing *E. coli* (STEC) overall and for *E. coli* O157:H7, specifically, as well 16 *Salmonella* serotypes. Values above 0 (the dotted line) indicate that the new model fit the data better than the previous model; values below 0 would indicate the opposite. No values fell below 0. *Differences in R^2^ were multiplied by 100.

The new Bayesian model builds on the previous frequentist model; both are negative binomial models with a log link function and a log offset for the population of each site each year. The previous frequentist model estimated illness count as a function of a categorical year variable and a second categorical variable, called site-entry year, that represented the year a given county in a given site started FoodNet surveillance (*1*). A modified Bayesian version of the previous frequentist model was implemented using an improper flat prior for the fixed effects and a fixed effect of site (instead of site-entry year) to enable comparison between the previous frequentist model and the new Bayesian model described here (Figures 1 & 2).

In the new model, illness count is modeled as a function of site and year, which is treated as a continuous variable. The new model allows for the estimation of separate, site-specific trends using penalized thin plate regression splines. These types of splines were selected because they provide computationally efficient, stable, and optimal low-rank approximations compared with thin plate splines, (ii) avoid issues related to knot placement through truncated eigen-decompositions, and (iii) allow model selection using methods dependent on model nesting (*2*). The priors and the value of the basis function used to represent the smooth term for the thin plate regression splines were set to the package default. For the fixed effects, an improper flat prior was used. For the splines, a half student-t prior was used with the scale parameter dependent on the standard deviation of the transformed response.

The new model was implemented with 6 chains and 10,001 iterations using the brms package (version 2.14.0) in R (version 3.6.2). Splines were implemented in the brms model using the s() function with the by parameter set to site so that separate site-specific splines were fit for each of the 10 sites. The new model generated a posterior predictive distribution of estimated mean log illness counts for each site in each year. We obtained samples from this distribution using the add_linpred_draws function in the tidybayes package (version 2.1.1) and exponentiated them. Illness estimates across sites were then summed for each draw-year combination to generate a distribution of illness estimates for the entire FoodNet catchment population for each year during the study period. Median incidence estimates and equal-tailed 95% credible intervals (CrI) were calculated for each year using these catchment-level distributions. The median and 95% CrIs were converted to incidence per 100,000 using the FoodNet catchment population for the specified year. To determine if incidence increased, decreased, or stayed the same, incidence estimates for the most recent year were compared with one or more reference year periods. For example, in the MMWR report published concomitantly with this paper, incidence estimates for 2021 were compared with average incidence estimates for 2016–2018. The incidence estimates for 2021 and the reference period were then used to calculate the percentage change for 2021 compared with the reference period.

## Data Availability

All data requests should be submitted to the Foodborne Diseases Active Surveillance Network, Enteric Disease Epidemiology Branch, Division of Foodborne, Waterborne and Environmental Disease, National Center for Emerging and Zoonotic Infectious Diseases, US Centers for Disease Control and Prevention.

## Ethics Statement

This activity was reviewed by US Centers for Disease Control and Prevention, National Center for Emerging and Zoonotic Infectious Diseases’ human subjects advisor, who determined that this effort was non-human-subjects research and exempt from IRB review.

